# A soluble bi-specific fusion protein for the improved expansion of human CD8^+^ CAR-T cells

**DOI:** 10.64898/2026.06.16.26355813

**Authors:** Jaclyn C. Law, Esther I. Matus, Pooja R. Mina, Amanda Sparkes, Navin Asokumar, Shannon Trottier, Gloria B. Kim, Jean Gariépy

## Abstract

The success of Chimeric Antigen Receptor (CAR) T cell therapy is heavily dependent on the quality of the final cellular product. Current expansion protocols often rely on reagents that require removal from cell culture media, posing logistical challenges in manufacturing, and can also lead to terminal differentiation. Here, we evaluate the use of a soluble, bead-free T cell activator, T cell expansion protein (T-CEP), as a streamlined alternative for generating potent CAR-T cells. Human T cells were activated with T-CEP or known T cell activators (Dynabeads and TransAct) and transduced with either CD19 or interleukin-13 (IL-13) mutein (tetravariant-13; TV-13)-based CAR lentiviral vectors. Our results demonstrate that T-CEP supports robust CAR-T cell expansion and achieves transduction efficiencies comparable to commercial reagents for both types of CAR-T cells. Notably, T-CEP significantly favored the expansion of CD8^+^ T cells, yielding an enhanced CD27^+^ phenotype and a lower CD4:CD8 ratio compared to TransAct. Cytotoxicity assays confirmed that T-CEP-expanded CAR-T cells possess cytolytic function equivalent to commercial reagents for both CARs, while exhibiting lower levels of inflammatory cytokine secretion. In summary, T-CEP represents a competitive alternative to existing expansion agents, as it does not require its removal during CAR-T manufacturing and generates a CD8^+^ dominant, less-differentiated phenotype without compromising efficacy.

**Key Points:**

- T-CEP is a soluble, bead-free activator that streamlines CAR-T manufacturing by eliminating the need for reagent removal from cultures.
- T-CEP consistently yields a desirable CD27^+^CD8^+^ phenotype with a moderate inflammatory profile compared to industry-standard expanders.

## Introduction

Resistance and disease relapse remain significant hurdles to the clinical efficacy of chimeric antigen receptor (CAR) T cell therapy in haematological malignancies.^1,2^ The quality of the infused CAR-T cell product is a crucial correlate of response but is limited by inconsistent cell function and phenotype.^2–5^ A low ratio of CD4:CD8 T cells, as well as a less differentiated phenotype, have been consistently associated with durable remission.^3,5–9^ However, achieving a product enriched for these phenotypes is dependent on the manufacturing process, where the outcome of T cell activation is governed by stimulatory signal strength and duration, as well as the mechanical context in which they are delivered.^10–14^ The consistent generation of CAR-T cells with defined phenotypes therefore remains a critical challenge.

CAR-T cells are manufactured by activating autologous T cells *ex vivo*, transducing them with a viral or non-viral CAR construct, followed by expansion.^15^ T cell activation requires T cell receptor (TCR) signaling, co-stimulation primarily though the CD28 co-receptor and cytokine signaling.^16^ In manufacturing, TCR and co-stimulatory signals are often triggered by agonistic monoclonal anti-human CD3 and CD28 antibodies immobilized on magnetic beads, such as Dynabeads (Thermofisher), or on polymeric scaffolds, such as human T cell TransAct (Miltenyi Biotec).^15^ Cytokines, commonly interleukin-2 (IL-2), are supplemented in the media.^15^ Despite the common application of these activators in the clinic, they frequently yield terminally differentiated cells, variable CD4:CD8 ratios and require additional de-beading or matrix removal steps during manufacturing.^14,15,17,18^

In our previous study, we developed a tandem single-chain variable fragment (scFv) bispecific T cell expansion protein (T-CEP) that delivers CD3 and CD28 signals in a soluble format.^19^ T-CEP demonstrated superior expansion of less differentiated, predominantly CD8^+^ T cells compared to both soluble and immobilized antibody methods, highlighting the potential utility of T-CEP in cell therapy manufacturing.^19^ In this report, we investigated the use of T-CEP for the generation of CAR-T cells and compare the quality of the expanded cells against those generated from Dynabeads and TransAct. Using healthy donor-derived T cells, we assessed the impact of T-CEP on CAR lentivirus transduction efficiency and subsequent expansion kinetics, cellular composition, phenotype and functionality of the CAR-T cells. We further analyzed the use of T-CEP for the transduction of two different CAR constructs to establish the versatility of T-CEP. We utilize a CD19 CAR-T cell construct that is approved for the clinical treatment of haematological tumours, and a solid tumour-targeted IL-13 mutein (tetravariant-13; TV-13)-based CAR-T construct containing four amino acid substitutions engineered to enhance selectivity for IL-13Rα2 over IL-13Rα1, an antigen highly expressed on high-grade glioblastoma.^20–22^ We demonstrate that T-CEP yields CAR-T cell products of superior CD4:CD8 T cell ratios and subset differentiation, with comparable functionality to leading commercial manufacturing reagents.

## Methods

Expanded methods are available in supplementary material.

### T-CEP protein expression and purification

The bispecific protein T-CEP was produced as previously described.^19^ A synthetic DNA gene encoding an anti-human CD28-scFv linked to an anti-human CD3-scFv was cloned into a pcDNA-3.4 expression plasmid and used to transform Expi293F cells (ThermoFisher). The expressed and secreted T-CEP was purified using HisTrap HP columns (GE Healthcare), then desalted into PBS using PD-10 columns (Cytiva). The purified protein was passed through endotoxin removal columns and sterilized by filtration.

### PBMCs and T cell isolation

Healthy donor human peripheral blood mononuclear cells (PBMCs) were isolated from EDTA-treated whole blood samples using density gradient centrifugation (Ficoll-Paque Plus, GE Healthcare) and cryopreserved in heat-inactivated fetal bovine serum (HI-FBS, Wisent Inc.) with 10% DMSO (ThermoFisher) in liquid nitrogen. PBMCs were collected under the Institutional Sunnybrook Research Ethics Board approval (Project Identification #2978). CD3^+^ T cells were isolated from PBMCs using the EasySep™ Human T Cell Isolation Kit (STEMCELL Technologies) per manufacturer’s instructions.

### CAR-T cell generation

CD3^+^ T cells were plated in a 96-well flat-bottom plate (Corning Inc.) in T cell medium (X-VIVO 15, Lonza; 5% HI-FBS; 100 IU/ml recombinant human IL-2, STEMCELL Technologies) and stimulated with Dynabeads, TransAct, or T-CEP (10ng/ml). After 24 hours, 50% of spent medium was replaced with fresh T cell medium containing a lentiviral vector encoding a GFP-tagged CD19 CAR (BPS Bioscience) or GFP-tagged TV-13 CAR lentivirus at a MOI of 20:1. After 48 hours, Dynabeads or TransAct were removed by magnetic separation or washing, respectively. T cells were transferred to a G-Rex 24-well plate (Wilson Wolf) and medium was exchanged every three days through replacing 75% of the spent medium for 12-13 days. On day 12, CD19 CAR-T cells were sorted by fluorescence activated cell sorting (BD FACSAria) based on GFP expression and DAPI (Thermofisher) exclusion for live cells.

### TV-13 CAR Lentivirus generation

TV-13 was inserted into a second-generation CAR cassette containing a CD8α signal peptide, CD8α hinge and transmembrane domains, a 4-1BB costimulatory intracellular signaling domain, and a CD3ζ activation domain.^22^ TV-13 CAR lentiviral particles were generated as previously described.^21,22^ Briefly, the TV-13 CAR plasmid was combined with third-generation packaging plasmids pMDLg/pRRE, pRSV-Rev, and pMD2.G in Opti-MEM, then added dropwise to a separate mixture of Opti-MEM and Lipofectamine 2000 to generate the transfection medium. The transfection medium was added dropwise to confluent HEK293T cells. Lentivirus-containing supernatants were collected at 24 and 48 hours after transfection, filtered, and concentrated by ultracentrifugation. Aliquots of lentiviral particles were stored at −80°C.

### xCELLigence real time cell analysis (RTCA) cytotoxicity assay

The xCELLigence RTCA DP system (Agilent Technologies) was used to monitor target cell growth and killing by CAR-T cells. To assess killing by CD19 CAR-T cells, anti-CD40 antibodies were immobilized on E-Plate 16 using the xCELLigence anti-CD40 IMT assay tethering kit (Agilent Technologies), and CD19^+^ Daudi cells were seeded at 4×10^4^ cells per well in RPMI (Wisent Inc.; with 10% HI-FBS, 1% penicillin and streptomycin).^23^ To monitor TV-13 CAR-T cell cytolytic activity, IL-13Rα2-expressing target U87 cells were seeded at 5,000 cells per well on E-Plate 16 in cDMEM (Wisent Inc.; with 10% HI-FBS and 1% penicillin and streptomycin). Target cell growth was monitored for 24 hours. Then, non-adhered cells and medium were removed, and CAR-T cells were seeded at 5:1 effector-to-target ratio over adherent cells. Monitoring resumed for 40-46 hours.

For both CAR-T cell models, each independent experiment was performed using two donors, for a total of 5-6 donors. Cell index was recorded as electrical impedance caused by cell adherence using RTCA Software Pro (Agilent Technologies). Data are reported as normalized cell index calculated per manufacturer’s “Calculation Principles” protocol:

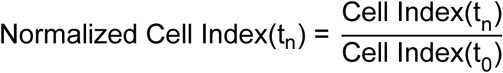

where t_n_ = Cell Index at a given time, and t_0_ = Cell Index just before the addition of CAR-T cells. Cytotoxic activity is reported as percent cytolysis:

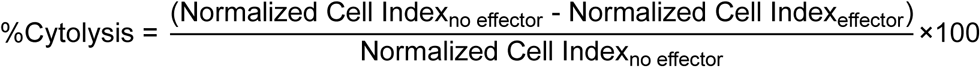

### Flow cytometry

Antibody clones and fluorophores are described in supplemental methods. CD19 CAR-T cells were incubated with Human TruStain FcX Fc blocker (BioLegend) for 10 minutes on ice, then stained with an antibody cocktail containing anti-human CD4, CD8, CD27 and CD45RA antibodies on ice for 30 minutes in HBSS (Wisent Inc.). Cells were washed with HBSS, then resuspended in DAPI.

To monitor the transduction and phenotype of TV-13 CAR-T cells, cells were incubated with Human TruStain FcX for 10 minutes at 4°C, then stained with eBioscience™ Fixable Viability Dye eFluor™ 450, anti-human CD4, CD8, CD27 and CD45RA in staining buffer (PBS with 2% HI-FBS) at 4°C for 30 minutes. To assess activation, T cells were re-stimulated with U87 cells for 46 hours, then washed with staining buffer. Cells were incubated with Human TruStain FcX for 10 minutes at 4°C, then incubated with eBioscience™ Fixable Viability Dye eFluor™ 450, anti-human CD8, CD4, CD27, CD45RA, CD25 or PD-1, and CD69 or LAG-3 for 30 minutes at 4°C. Cells were washed and resuspended in staining buffer, then analyzed on the BD FACSymphony A3 Flow Cytometer.

### Cytokine analysis

Following 46-hours co-culture with or without U87 target cells, cell culture supernatants were harvested, centrifuged and stored at −80°C until time of assay. The LEGENDplex™ Human CD8/NK mix and match kit (BioLegend) was used to measure the secretion of interferon-γ (IFN-γ), tumor necrosis factor-α (TNF-α), IL-4 and Granzyme B. The assay was performed per manufacturer’s instructions and samples were analyzed on the BD FACSymphony A3 Flow Cytometer.

### Data analysis

Flow cytometric analyses were performed using FlowJo v10 (BD Biosciences). Secreted cytokines were analyzed using BioLegend LEGENDplex™ Data Analysis Software Suite fitted with a 5-parameter logistic curve, then normalized by square root transformation. Statistical analyses were performed using GraphPad Prism 10. Results with a *p*-value <0.05 were considered significant.

## Results

### Human CD19 CAR-T cells expanded with T-CEP are predominantly CD8^+^ T cells

We first utilized CD19-directed CAR-T cells as a benchmarking model.^20^ The expansion of CD19 CAR-T cells by T-CEP was compared to those expanded with the industry-standard activators: Dynabeads or TransAct. The final CAR-T cell yield was comparable between the 3 different expanders tested, ranging from an average of 8×10^6^ to 1×10^7^ CAR-T cells from five healthy donors from an initial seeding density of 2×10^5^ cells (Figure 1A). CD19 CAR lentivirus transduction efficiency was assessed in each T cell compartment on day 12 based on their GFP expression (Figure 1B). While the frequency of CAR^+^ CD4^+^ T cells was comparable between the three expanders, there was a significantly lower frequency of CAR^+^ CD8^+^ T cells in the TransAct pool (Figure 1B).

**Figure 1.**
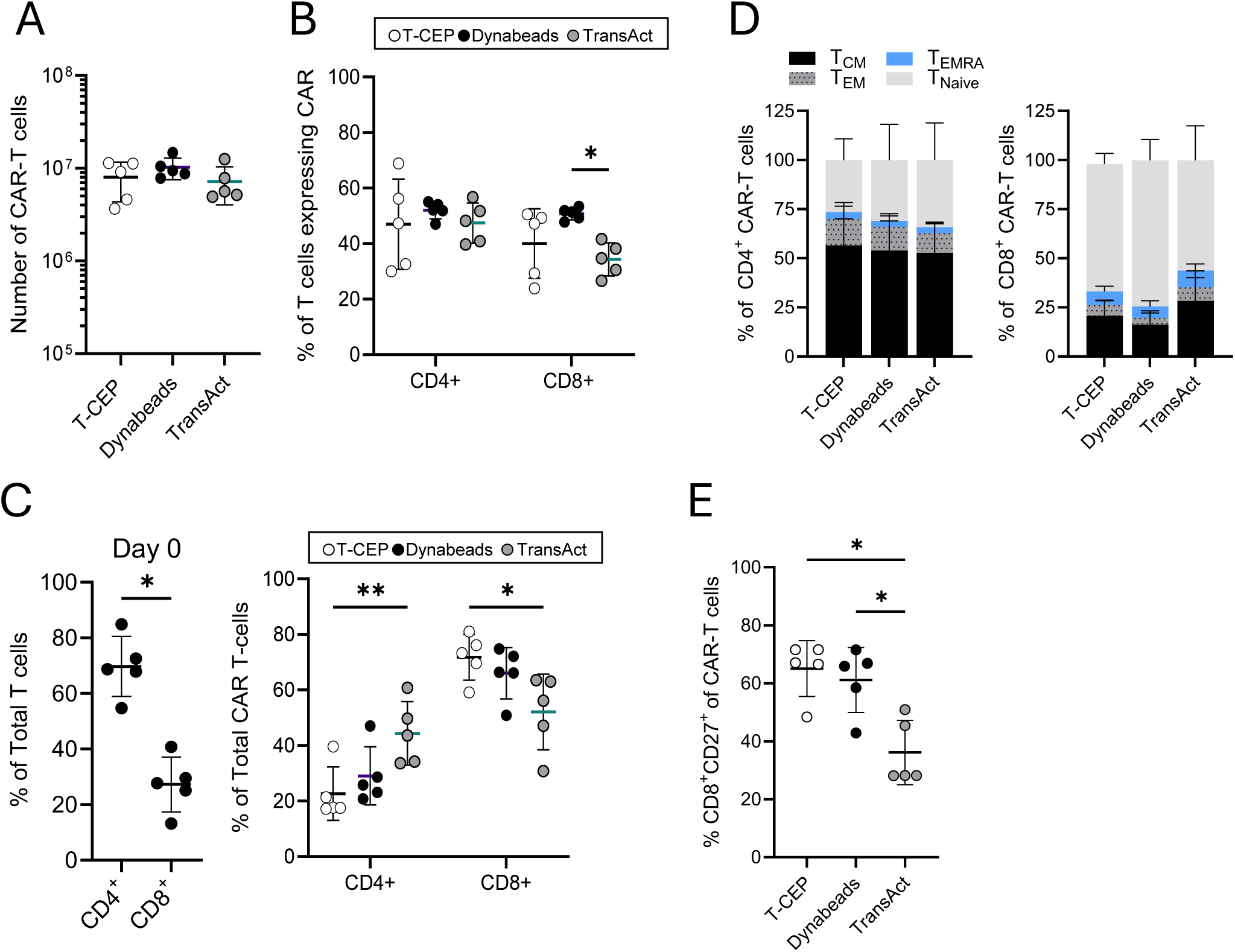
Phenotypic analysis of human CD19-CAR-T cells expanded for 12 days. T cells were harvested from GREX-24 well plates after 12 days of expansion with different activators. **(A)** Total number of CAR-T cells after 12 days of expansion. **(B)** The transduction efficiency of donor T cells using GFP-expressing CD19-CAR lentivirus with different activators. **(C)** The frequency of CD4^+^ and CD8^+^ T cells on Day 0 (left) and on Day 12 within the transduced CAR-T cells (right). **(D)** The expression of CD45RA and CD27 on CD4^+^ (left) and CD8^+^ (right) human CAR-T cell populations as an indicator of differentiation, where T_EMRA_ = CD45RA^+^CD27^-^, T_EFF_ = CD45RA^-^CD27^-^, T_CM_ = CD45RA^-^CD27^+^ and T_Naive_ = CD45RA^+^CD27^+^. **(E)** Proportion of CD8^+^ CAR-T cells expressing CD27. Graphs show mean±SD, n=5 donors. Statistical analyses were performed using RM 2-way ANOVA, Tukey’s multiple comparisons test. \**p<0.05*, \*\**p<0.01*.

We previously showed that T-CEP activation preferentially expands CD8^+^ T cells within polyclonal populations.^19^ However, it remained unclear whether preferential CD8^+^ expansion could be consistently achieved following CAR transduction. Thus, we evaluated the frequency of CD4^+^ and CD8^+^ subsets within the final CAR-T cell products from each expander (Figure 1C). All five donors used in the study started with a higher proportion of CD4^+^ T cells, averaging 68% CD4^+^ and 27% CD8^+^ T cells (mean CD4:CD8 ratio of 2.5) (Figure 1C, left). By day 12, T-CEP significantly favored CD8^+^ T cell expansion, yielding the highest frequency of CD8^+^ cells (71%) and a markedly lower CD4:CD8 ratio (0.32) compared to TransAct (52% and 0.85, respectively; Figure 1C). These results demonstrate that T-CEP-mediated activation skews the final CAR-T product toward a predominantly CD8^+^ population.

We next evaluated the memory phenotype of the expanded cells. We previously observed that T-CEP enhanced CD27 expression.^19^ Importantly, CAR-T cells derived from the CD27-expressing central memory (T_CM_) or naïve (T_Naive_) subsets have been linked to improved clinical outcomes and CAR-T cell persistence *in vivo*.^6,24^ Here, we show that the distribution of each memory subset, defined by the expression of CD45RA and CD27, is comparable between the different T cell activators on day 12 within both CD4^+^ and CD8^+^ populations (Figure 1D). However, CD27 expression, representing the combined proportions of less-differentiated T_CM_ and T_Naive_, was significantly enriched on CD8^+^ CAR-T cells expanded with T-CEP or Dynabeads when compared to TransAct, consistent with our previous observations (Figure 1E).^19^ CD27 expression on CD4^+^ CAR-T cells was similar between the three expanders (data not shown). Altogether, T-CEP activation enriches for desirably less differentiated, predominantly CD8^+^ CAR-T cells compared to TransAct.

### CD19 CAR-T cells expanded with T-CEP demonstrate potent cytotoxicity *in vitro*

To assess the cytotoxic capacity of the CAR-T cells expanded with T-CEP, Dynabeads or TransAct, CD19 CAR-T cells were co-cultured with immobilized CD19^+^ Daudi cells at a 5:1 effector-to-target cell ratio in the xCELLigence RTCA system.^23^ Cytotoxicity was monitored based on cell index over time. In negative control wells containing only Daudi cells, and wells containing untransduced T cells, the cell index continues to rise (Figure 2A). All other wells containing both Daudi cells and CAR-T cells show decreasing cell index over time, indicating Daudi cell death (Figure 2A). Significant killing by CAR-T cells, expressed as percent cytolysis, is observed by 6 hours after co-culture (Figure 2B). By 40 hours of co-culture, 84% cytolysis is achieved by T-CEP expanded CAR-T cells, compared to 64% and 61% cytolysis by Dynabeads and TransAct expanded CAR-T cells, respectively (Figure 2B). However, this difference did not reach statistical significance (Figure 2B).

**Figure 2.**
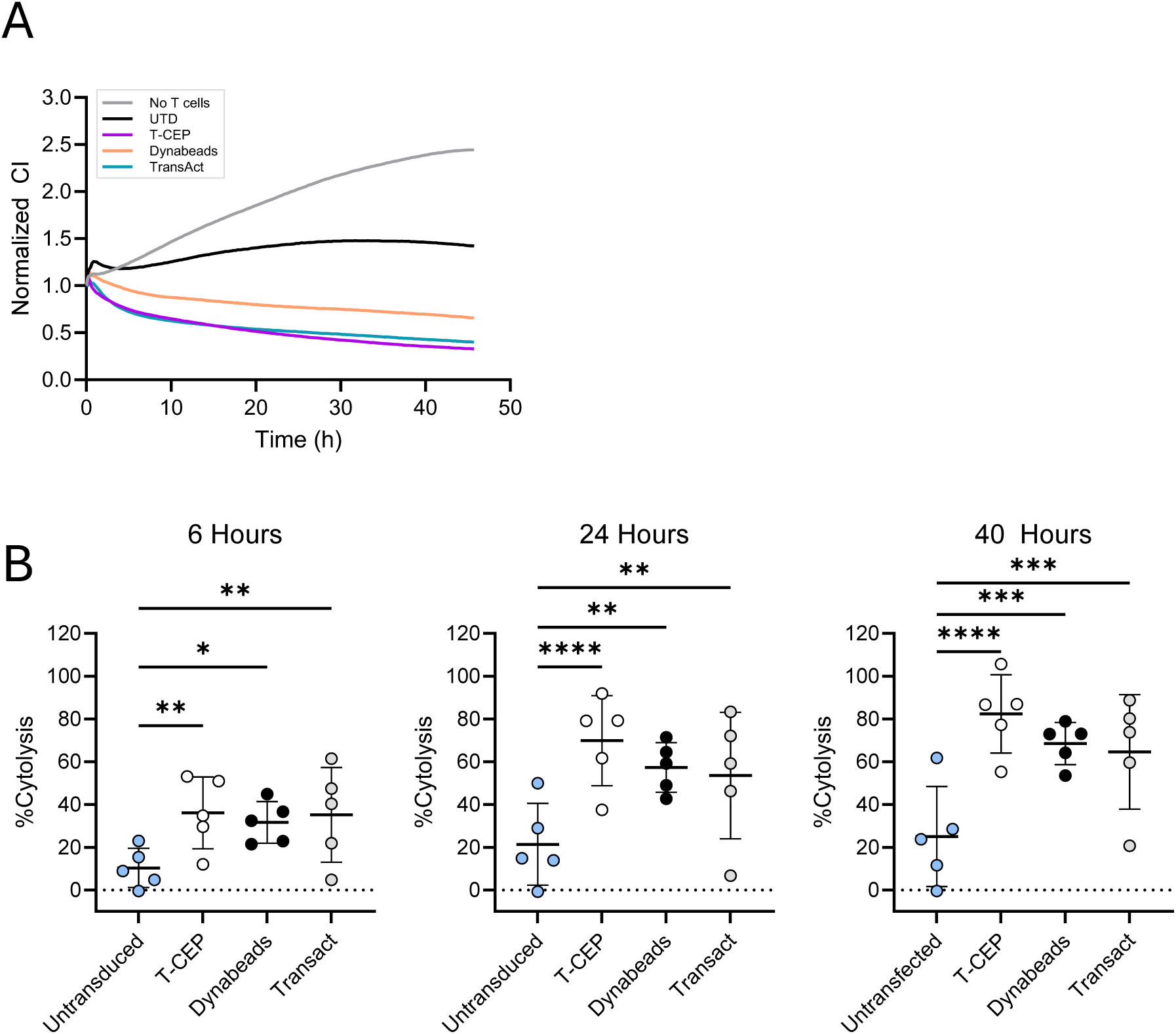
Cytotoxicity of CD19 CAR-T cells against CD19-expressing Daudi cells. On Day 12 of expansion, CAR-T cells were harvested and seeded onto CD19^+^ Daudi cells in the xCELLigence RTCA system. Cell index was recorded for up to 46h. **(A)** Normalized cell index (CI) of CD19^+^ Daudi cells over time when incubated with or without CAR-T cells expanded with the indicated reagent. Plot shows the mean CI of one RTCA experiment (n=2). The experiment was repeated twice for a total of n=5. UTD = untransduced T cells. **(B)** Graphs depict the percent cytolysis of CD19^+^ Daudi cells 6h, 24h and 40h after co-incubation with CD19 CAR-T cells expanded with each reagent. Graphs show mean±SD, n=5 donors. Statistical analyses were performed using 1-way RM ANOVA, Tukey’s multiple-comparisons test. *\* p<0.05, ** p<0.01, *** p<0.001, **** p<0.0001*.

### TV-13 CAR-T cells expanded with T-CEP are predominantly CD27^+^CD8^+^ T cells

To validate T-CEP as a broadly applicable platform, we utilized the IL-13Rα2-targeted TV-13 CAR to confirm that the previously observed expansion kinetics and memory phenotypes are maintained independent of CAR specificity. T cells were expanded with T-CEP, Dynabeads or TransAct for 13 days (Figure 3A). The total T cell yield was comparable between the different expanders, achieving a median of 3.0×10^7^ total cells by day 13 (Figure 3B). Across all three expanders, TV-13 CAR-T cells, identified by GFP-expression, exhibited similar expansion kinetics, peaking at day 9 followed by a moderate decline by day 13 (Figure 3C). Transduction in this model across all expanders favored CD4^+^ over CD8^+^ T cells, achieving a mean of 83% GFP^+^ and 49% GFP^+^, respectively (Figure 3D). Dynabeads activation led to significantly greater CD4^+^ transduction compared to T-CEP and TransAct (Figure 3D). Despite more efficient transduction in CD4^+^ T cells, expansion kinetics reveal a notably reduced CD4:CD8 ratio in total T cells by day 13 in T-CEP expanded cultures (mean ratio 0.2) compared to Dynabeads (0.6) and TransAct (0.3) (Figure 3E). Importantly, this skew toward CD8^+^ T cells is reflected within the final CAR-T cell product. An average of 76% of CAR-T cells were CD8^+^ when expanded with T-CEP, compared to 52% and 62% after expansion with Dynabeads and TransAct, respectively (Figure 3F).

**Figure 3.**
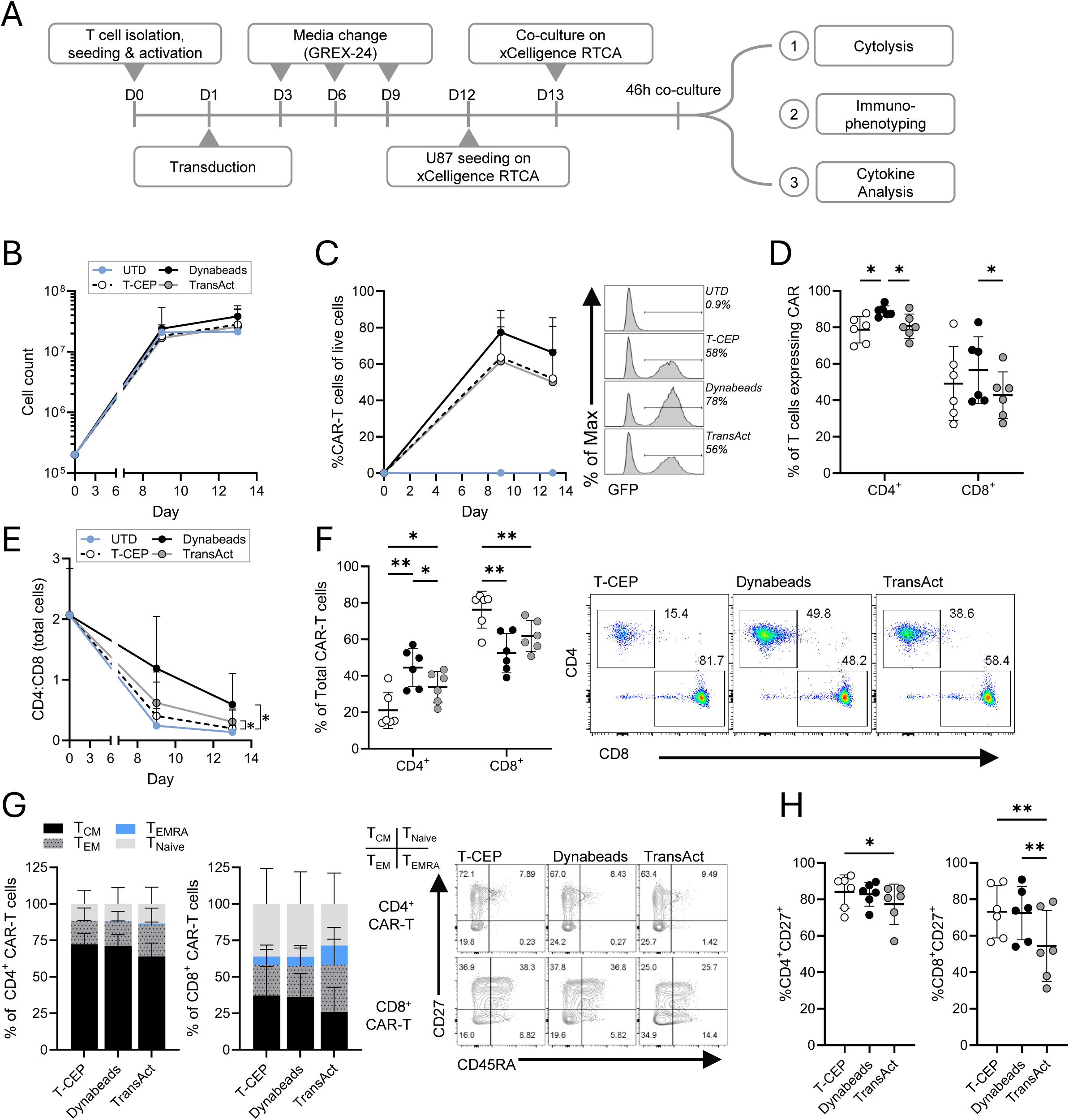
Expansion of TV-13 CAR-T cells using different T cell expanders. T cells were harvested from Grex-24 well plates after 13 days of expansion with different activators. **(A)** Experimental workflow for CAR-T cell expansion and downstream assays. **(B)** Total number of T cells expanded, UTD = Untransduced T cells. **(C)** The frequency of total GFP^+^ CAR-T cells on Days 9 and 13 of expansion with different activators. Histograms show the frequency of GFP^+^ live cells from 1 donor on Day 13. **(D)** The transduction efficiency of GFP-tagged TV-13-CAR lentivirus in CD4^+^ and CD8^+^ T cells. **(E)** The ratio of total CD4^+^:CD8^+^ T cells throughout expansion with different activators. **(F)** CD4^+^ and CD8^+^ populations within transduced T cells on D13 (left) and representative flow plots from one donor showing the frequency of CD4^+^ and CD8^+^ CAR-expressing T cells on D13 (right). **(G)** Memory distribution of expanded CD4^+^ and CD8^+^ CAR-T cells based on CD45RA and CD27 expression. Representative flow plots show the expression of CD45RA and CD27 by CD4^+^ and CD8^+^ CAR-T cells from 1 donor. **(H)** Proportion of CD27^+^ expressing CAR-T cells. All graphs show mean±SD, n=6 donors. Statistical analyses were performed using RM 2-way ANOVA, Tukey’s multiple comparisons test. \**p<0.05*, \*\**p<0.01*.

Since we observed higher CD27 expression on CD8^+^ T cells in the CD19 CAR-T cell model, we next analyzed the memory subsets of TV-13 CAR-T cells. Analysis of memory subsets based on CD27 and CD45RA expression revealed comparable memory subset distributions across all activation methods by day 13 for both CD4^+^ and CD8^+^ CAR-T cells (Figure 3G). However, both CD4^+^ and CD8^+^ CAR-T cells expanded with T-CEP contained a significantly higher frequency of CD27^+^ cells compared to those expanded with TransAct (Figure 3H). These data suggest that T-CEP favors the expansion of less differentiated CD8^+^ CAR-T cells independent of CAR-specificity.

### T-CEP generated TV-13 CAR-T cells maintain high cytolytic activity

To evaluate TV-13 CAR-T cell function, TV-13 CAR-T cells were co-cultured with U87 cells at a 5:1 effector-to-target cell ratio and cell index was monitored for 46 hours using the xCELLigence RTCA assay. The cell index remained constant in wells containing U87 alone or with untransduced T cells (Figure 4A). The cell index declined over time when U87 cells were co-cultured with TV-13 CAR-T cells (Figure 4A). The decline in cell index, expressed as percent cytolysis, was followed for 46 hours (Figure 4B). Significant cytolysis of U87 cells was observed by 12 hours of co-culture with CAR-T cells generated from T-CEP and Dynabeads but not TransAct (Figure 4C). By 24 hours of co-culture, there was significant cytolysis of U87 by CAR-T cells generated from all expanders (Figure 4C). By 46 hours, at the end of co-culture, 80% of U87 cells were lysed by T-CEP generated CAR-T cells, compared to 81% by Dynabeads and 71% by TransAct generated CAR-T cells (Figure 4C). Like our observations with CD19 CAR-T cells, TransAct-generated CAR-T cells achieved the lowest percent cytolysis, although this did not reach statistical significance in either model (Figures 4C, 2B). In parallel, TV-13 CAR-T cells co-cultured with U87 target cells at a 10:1 effector-to-target ratio reached levels of cytolysis by 46 hours comparable to those observed in the 5:1 co-culture (Figure S1). Again, at 10:1, TransAct-expanded CAR-T cells achieved the lowest percent cytolysis (65%) compared to T-CEP (72%) and Dynabeads (87%) expanded cells, although this difference was not statistically significant (Figure S1). Furthermore, cursory microscopic observation of T cell clusters after re-stimulation with U87 showed indistinguishable morphology between the different expanders (Figure 4D). These results indicate that T-CEP activation can generate cells of equivalent cytolytic potency and morphology as commercial activators, positioning T-CEP as a viable alternative for CAR-T cell expansion.

**Figure 4.**
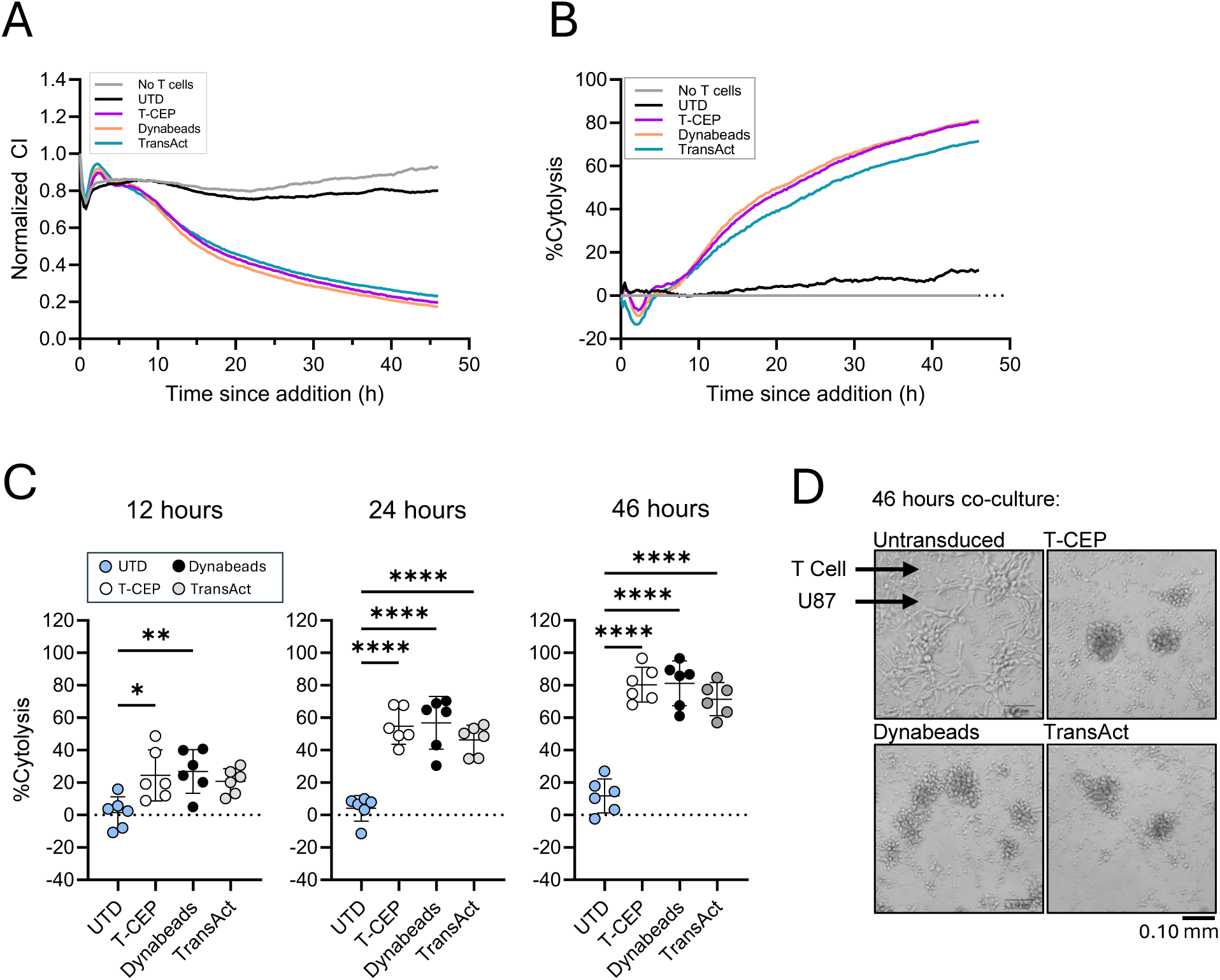
Cytotoxicity of TV-13 CAR-T cells against IL-13Rα2-expressing U87 cells. TV-13 CAR-T cells were harvested at the end of expansion and seeded onto IL-13Rα2^+^ U87 cells in the xCELLigence RTCA system. Cell index was recorded for up to 46h and CAR-T cells were harvested from the RTCA system for phenotyping. **(A)** Normalized cell index (CI) of IL-13Rα2^+^ U87 cells over time when incubated with or without CAR-T cells that were expanded with the indicated reagent. UTD = untransduced T cells. **(B)** Percent cytolysis of U87 cells over the entire course of co-culture. **(C)** Statistical analysis of the percent cytolysis of U87 cells at 12h, 24h and 46h co-culture with CAR-T cells. **(D)** 100X microscopic image of U87 cells after 46h co-culture with CAR-T cells from 1 donor. All graphs show n=6 donors. **(A)-(B)** show mean, **(C)** shows mean±SD. Statistical analyses were performed using 2-way RM ANOVA, Tukey’s multiple-comparisons test. \**p<0.05*, \*\**p<0.01, **** p<0.0001*.

### TV-13 CAR-T cells expanded with T-CEP exhibit robust cytokine expression with minimal exhaustion

Dynabeads and TransAct were removed on day 3 of cell culture by magnetic separation or a wash step, respectively. In contrast, T-CEP is maintained in the culture and diluted by subsequent partial media exchanges. To evaluate whether prolonged T-CEP exposure affects the final cell product, we analyzed the activation, exhaustion and cytokine profiles of TV-13 CAR-T cells. Following a 46-hour co-culture with or without U87 targets, we utilized flow cytometry to assess surface marker expression and cytokine secretion in the supernatant (Figure 5A, Figure S2). Without antigen stimulation, both CD4^+^ and CD8^+^ CAR-T cells exhibited minimal baseline expression of activation (<1% CD69^+^CD25^+^) and exhaustion (<0.5% PD-1^+^LAG-3^+^) markers (Figure 5B-C). These markers were significantly and comparably upregulated across all three expansion conditions only upon antigen encounter (Figure 5B-C). Importantly, the consistent lack of marker expression at baseline across all groups suggests that the continuous presence of T-CEP in the culture does not induce premature activation or exhaustion.

**Figure 5.**
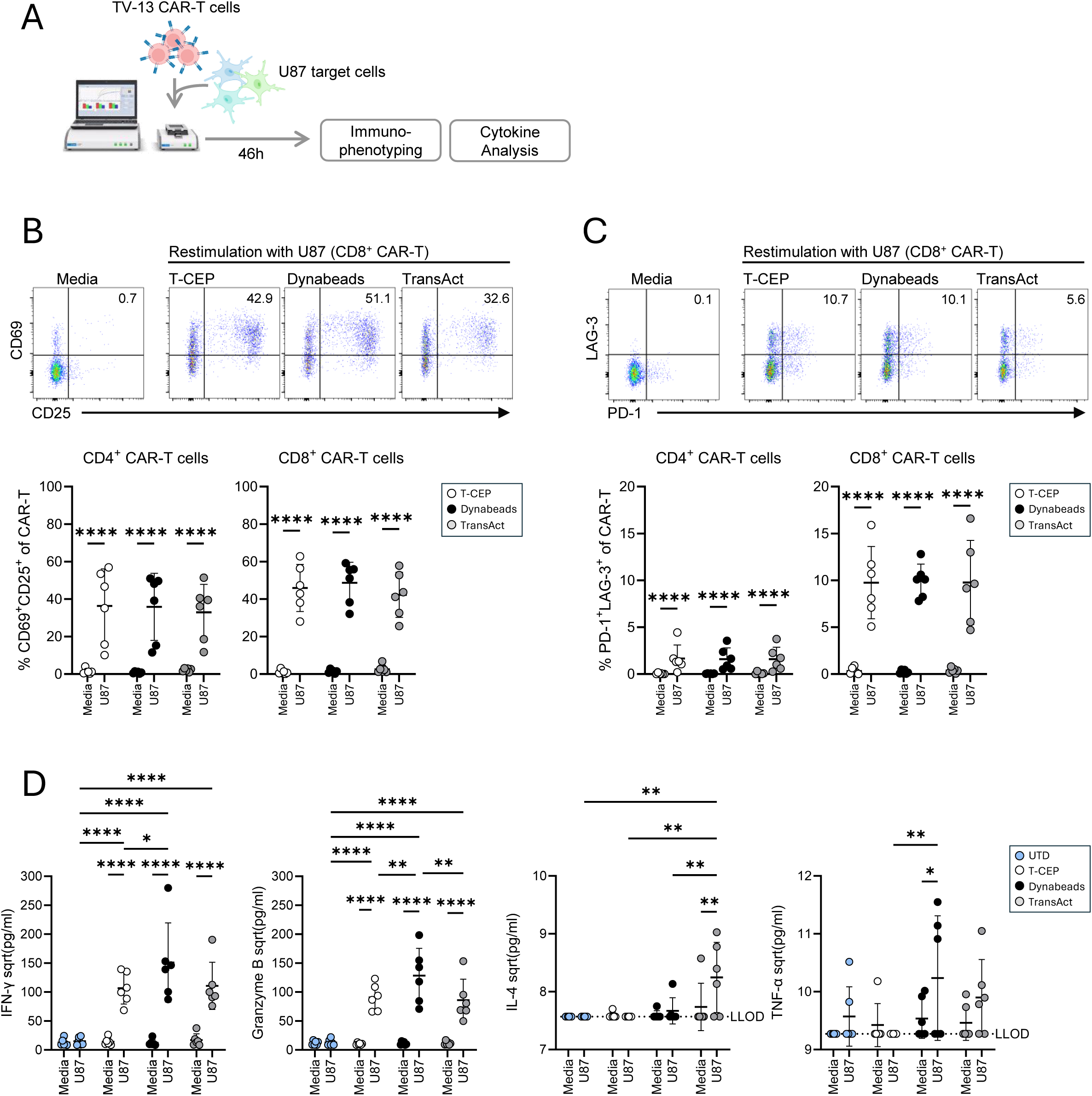
Phenotypic analysis of CAR-T cell responses to target cells. CAR-T cell surface markers and cytokine secretion were analyzed after 46h co-culture with IL-13Rα2-expressing U87 target cells at a 5:1 ratio. **(A)** Assay set up. **(B)** Graphs show co-expression of CD69 and CD25 activation markers by CD4^+^ and CD8^+^ CAR-T cells. Representative flow plots from 1 donor depict CD69 and CD25 expression by CD8^+^ CAR-T cells. **(C)** Graphs show co-expression of PD-1 and LAG-3 by CD4^+^ and CD8^+^ CAR-T cells. Representative flow plots from 1 donor depict PD-1 and LAG-3 expression by CD8^+^ CAR-T cells. **(D)** Secretion of IFN-γ, Granzyme B, IL-4 and TNF-α in cell culture supernatants by CAR-T cells after restimulation with U87 cells or with media as negative controls. UTD = untransduced T cells. LLOD = lower limit of detection. Graphs show mean±SD, n=6 donors. Icons in **(A)** are sourced from biorender.com. Statistical analyses were performed using 2-way RM ANOVA, Tukey’s multiple-comparisons test. \**p<0.05*, \*\**p<0.01, *** p<0.001, **** p<0.0001*.

We next examined the cytokine secretion profile of the CAR-T cell products. We show that IFN-γ and Granzyme B were highly expressed in response to antigen challenge by all CAR-T products (Figure 5D). Interestingly, Dynabeads-expanded CAR-T cells expressed significantly higher levels of IFN-γ and Granzyme B than CAR-T cells expanded with T-CEP or TransAct (Figure 5D). T-CEP and Dynabeads expanded cells did not secrete IL-4, indicating a type I response to antigen challenge (Figure 5D). IL-4 was significantly upregulated only by cells expanded with TransAct, albeit at a low level (Figure 5D). While low levels of the proinflammatory cytokine TNF-α were detected in most cultures, Dynabeads expanded cells were the only group to reach statistically significant levels of expression (Figure 5D). Altogether, these data indicate that while surface exhaustion and activation markers are consistent across conditions, variations in cytokine expression may suggest subtle functional distinctions between the resulting products.

## Discussion

While CAR-T cell therapy has made great strides in the treatment of haematological cancers, many patients are non-responsive, or relapse after treatment.^1,2^ Although patient- and tumor-specific factors may play a role in determining long-term remission, such as the patient’s age and indication, the quality of the therapeutic CAR-T cell product has emerged as a crucial correlate of response.^2–5,25^ In this study, we compared the performance of three anti-human CD3/CD28 based compounds for generating CAR-T cells of different specificities: a linear bispecific protein, T-CEP, and two clinically established activators, Dynabeads and TransAct. T-CEP not only consistently achieved robust CD8^+^ expansion across donors and varying CAR architecture (targeting hematological and solid tumours), but also produced cells of comparable cytolytic activity and activation.

The significance of product composition on anti-tumor effects is well-established. Clinical trials demonstrate that a 1:1 CD4:CD8 ratio yields clinical responses superior to unselected CAR-T products. ^26,27^ The impact of CD8^+^ CAR-T enrichment has not been directly explored in the clinic, although preclinical mouse models suggest there may be an effective therapeutic range between 3:1 to 1:3. ^6,28^ Nevertheless, low CD4:CD8 ratios have been identified as a prognostic indicator of CAR-T efficacy.^7^ Indeed, disease remission and survival for CD19-haematological cancers, and more recently for Il-13Rα2-targetted high-grade glioma, have been consistently associated with elevated CD27^+^CD8^+^ CAR-T cells across several clinical trials.^3,5,6,8,9^ CD27 co-stimulation promotes CAR-T cell survival, expansion and memory differentiation.^29^ Reduced central and stem-like memory cells were instead associated with disease relapse.^30^ In fact, combinatorial infusion products comprising CD8^+^ T_CM_ and CD4^+^ T_Naive_/T_CM_ CAR-T subsets yielded superior anti-tumor reactivity.^6^ Recent engineering strategies have further sought to integrate CD27-signaling domains directly into the CAR construct to optimize anti-tumor responses.^29,31^ The capacity of T-CEP to consistently generate a CD27^+^-enriched CD8^+^ T cell population aligns with these clinical correlates.

Accordingly, a primary challenge in CAR-T cell production is the generation of sufficient effector CD8^+^ CAR-T cells of a desirable memory phenotype despite variability in the donor starting material.^2^ Commonly used expansion platforms, including Dynabeads and TransAct, produce variable CD4:CD8 T cell ratios.^17,18^ 4-1BB-CD19 CAR-T products are frequently dominated by up to 80% CD4^+^ T cells.^1^ However, expanding CD8^+^ CAR-T cells in the absence of CD4^+^ T cells significantly reduces its anti-tumor activity and persistence *in vivo*, establishing a crucial role for CD4^+^ T cell help in producing CD8^+^ effectors.^32^ The requirement for isolation steps to achieve ideal CD4:CD8 ratios of specific memory subsets introduces significant complexity into the manufacturing process. T-CEP addresses these issues by consistently promoting CD8 expansion across donors with various starting T cell distributions and across different CAR architectures without needing manual enrichment. Crucially, T-CEP favours the expansion of less differentiated CD8^+^ T cells without depleting the CD4^+^ compartment, thereby preserving T-helper signals necessary for the long-term function and persistence of CD8^+^ CAR-T cells. In addition, we demonstrate two CAR lentiviruses with distinct transduction patterns: the CD19-CAR transduces CD4^+^ and CD8^+^ T cells equally, while the TV-13-CAR shows a significant transduction preference for CD4^+^ cells (Figures 1B, 3D). The T-CEP platform can overcome this imbalanced transduction of CD4^+^ T-helper cells by promoting the robust expansion of CD8^+^ cells (Figure 3F).

We also observed a reduction in IFN-γ and Granzyme B secretion, and no TNF-α, by T-CEP-expanded TV-13 CAR-T cells compared to Dynabeads, suggesting that there may be a distinct functional pattern that maintains robust cytolytic activity with reduced inflammatory signaling. In the clinical application of CAR-T therapy, cytokine release syndrome occurs in 30–100% of patients shortly after infusion, driven largely by systemic elevation of inflammatory mediators such as IFN-γ and TNF-α.^33^ Given that blocking IFN-γ remediates these toxicities while preserving cytolytic activity, the capacity to reduce the inflammatory output of the infusion product may be desirable.^34^

In our study, this attenuated cytokine profile is not likely indicative of cellular exhaustion or chronic stimulation (Figure 5D). Although T-CEP remains in the culture throughout expansion, while Dynabeads and TransAct are removed on day 3, the final product maintains a less-differentiated phenotype with killing capacity identical to Dynabeads and TransAct. If the sustained presence of T-CEP were driving exhaustion, we would expect a significant decline in cytolysis and a shift toward a more differentiated effector state, as well as upregulation of exhaustion markers at baseline.^35^ It is unlikely that the cytokine signature observed in T-CEP expanded cells is a consequence of preferential expansion of CD8^+^ T cells. In this study, we show that TransAct generates a ratio of CD4:CD8 CAR-T cells comparable to Dynabeads, yet TransAct consistently yielded lower cytokine levels that aligned more closely with the T-CEP profile than Dynabeads (Figures 1C, 3F, 4D). The current data suggest that the selected activation platform may alter the functional pathways driving cytotoxicity.

While the precise biophysical mechanisms driving preferential CD8^+^ expansion were not the primary focus of this study, our findings align with evidence suggesting that the physical state of the activation signal serves as a critical determinant of lineage bias. T cell activation is affected by the membrane fluidity of antigen presenting cells.^36,37^ Specifically, T cells activated on flexible, low-stiffness surfaces demonstrate a natural preference for CD8^+^ expansion over those stimulated by rigid substrates.^13^ Expansion platforms such as Dynabeads and TransAct rely on rigid immobilization of anti-CD3 and anti-CD28 full-length antibodies onto superparamagnetic beads or nanomatrices, respectively.^15,38^ This immobilization strategy differs fundamentally from the soluble presentation utilized by T-CEP. The observation that soluble delivery favours CD8^+^ enrichment is further supported by recent reports of a nanocage system, which similarly leverage soluble anti-CD3 and anti-CD28 to bias T cell compositions toward a CD8^+^ effector phenotype (Nanotein Technologies).^39^

Finally, while we provide compelling evidence for the use of T-CEP in CAR-T cell manufacturing using healthy donor cells, T-CEP’s performance remains to be validated using patient-derived starting material. The physiological impact of tumor burden and prior exposure to standard-of-care therapies may significantly impact the baseline exhaustion and activation states of the starting T cells, as well as the final product.^2,40^ Furthermore, the long-term therapeutic potential of T-CEP-expanded cells remains to be validated *in vivo*, such as the influence of a CD8-skewed product on sustained anti-tumor immunity, and its ability to form durable memory during tumour rechallenge. Nevertheless, given that the CAR-T cells generated here are functionally comparable to those produced by industry-standard expanders, we anticipate similar, if not superior, performance *in vivo*. As such, our data highlight T-CEP as a viable and highly efficient tool for the generation of potent, phenotypically robust CAR-T cell products.

## Data Availability

All data produced in the present study are available upon reasonable request to the corresponding authors.

## Acknowledgements

We thank Dr. Hon Leong for providing U87 cells, Dr. Kevin Conrad for assistance with flow cytometry, the healthcare team at the Sunnybrook Health Sciences Centre Transfusion Medicine Clinic for assistance with sample collection, and all the blood donors for participating to make this study possible. This work was supported by the Canadian Institutes of Health Research (CIHR) project grants PJT-468907 and PJT-517882 to JG.

## Authorship Contributions

JCL and EIM designed and performed experiments. JCL analyzed data and created the figures. JCL and JG wrote the manuscript. AS and NA provided technical assistance. JG and GBK conceptualized the study. GBK designed TV-13 lentivirus construct and developed the methodology for TV-13 CAR lentivirus generation. GBK, PRM and ST generated TV-13 lentivirus. PRM and GBK performed preliminary studies. JG and GBK provided scientific oversight. JCL, JG and GBK edited the manuscript.

## Data Sharing Statement

For original data, please contact

## References

1. Haradhvala NJ, Maus M V. Understanding Mechanisms of Response to CAR T-cell Therapy through Single-Cell Sequencing: Insights and Challenges. Blood Cancer Discov. 2024;5(2):86–89. doi:10.1158/2643-3230.BCD-23-0212

2. Baguet C, Larghero J, Mebarki M. Early predictive factors of failure in autologous CAR T-cell manufacturing and/or efficacy in hematologic malignancies. Blood Adv. American Society of Hematology. 2024;8(2):337–342. doi:10.1182/bloodadvances.2023011992

3. Haradhvala NJ, Leick MB, Maurer K, et al. Distinct cellular dynamics associated with response to CAR-T therapy for refractory B cell lymphoma. Nat Med. 2022;28(9):1848–1859. doi:10.1038/s41591-022-01959-0

4. Lin RJ, Lobaugh SM, Pennisi M, et al. Impact and safety of chimeric antigen receptor T-cell therapy in older, vulnerable patients with relapsed/refractory large B-cell lymphoma. Haematologica. Ferrata Storti Foundation. 2021;106(1):255–258. doi:10.3324/haematol.2019.243246

5. Fraietta JA, Lacey SF, Orlando EJ, et al. Determinants of response and resistance to CD19 chimeric antigen receptor (CAR) T cell therapy of chronic lymphocytic leukemia. Nat Med. 2018;24(5):563–571. doi:10.1038/s41591-018-0010-1

6. Sommermeyer D, Hudecek M, Kosasih PL, et al. Chimeric antigen receptor-modified T cells derived from defined CD8+ and CD4+ subsets confer superior antitumor reactivity in vivo. Leukemia. 2016;30(2):492–500. doi:10.1038/leu.2015.247

7. Galli E, Bellesi S, Pansini I, et al. The CD4/CD8 ratio of infused CD19-CAR-T is a prognostic factor for efficacy and toxicity. Br J Haematol. 2023;203(4):564–570. doi:10.1111/bjh.19117

8. Brown CE, Hibbard JC, Alizadeh D, et al. Locoregional delivery of IL-13Rα2-targeting CAR-T cells in recurrent high-grade glioma: a phase 1 trial. Nat Med. 2024;30(4):1001–1012. doi:10.1038/s41591-024-02875-1

9. Cheloni G, Karagkouni D, Pita-Juarez Y, et al. Durable response to CAR T is associated with elevated activation and clonotypic expansion of the cytotoxic native T cell repertoire. Nature Communications. 2025;16(1). doi:10.1038/s41467-025-59904-x

10. Bortoletto N, Scotet E, Myamoto Y, D’Oro U, Lanzavecchia A. Optimizing anti-CD3 affinity for effective T cell targeting against tumor cells. Eur J Immunol. 2002;32(11):3102–3107. doi:10.1002/1521-4141(200211)32:11<3102::AID-IMMU3102>3.0.CO;2-C

11. O’Connor RS, Hao X, Shen K, et al. Substrate rigidity regulates human T cell activation and proliferation. J Immunol. 2012;189(3):1330–1339. doi:10.4049/jimmunol.1102757

12. Judokusumo E, Tabdanov E, Kumari S, Dustin ML, Kam LC. Mechanosensing in T lymphocyte activation. Biophys J. 2012;102(2):L5–7. doi:10.1016/j.bpj.2011.12.011

13. Anandasivam N, Ali R, Gustinvil L, Rosenwasser MJ, Dunlop IE, Delcassian D. Tunable Enhancement of T Cell Expansion Through Modulation of Stiffness and Adhesion Receptor Engagement in an Engineered Hydrogel Platform. Advanced Materials. Published online February 20, 2026. doi:10.1002/adma.202505965

14. Zhang DKY, Adu-Berchie K, Iyer S, et al. Enhancing CAR-T cell functionality in a patient-specific manner. Nat Commun. 2023;14(1):506. doi:10.1038/s41467-023-36126-7

15. Wang X, Rivière I. Clinical manufacturing of CAR T cells: Foundation of a promising therapy. Mol Ther Oncolytics. Nature Publishing Group. 2016;3:16015. doi:10.1038/mto.2016.15

16. Smith-Garvin JE, Koretzky GA, Jordan MS. T cell activation. Annu Rev Immunol. 2009;27:591–619. doi:10.1146/annurev.immunol.021908.132706

17. Yang S, Dudley ME, Rosenberg SA, Morgan RA. A simplified method for the clinical-scale generation of central memory-like CD8+ T cells after transduction with lentiviral vectors encoding antitumor antigen T-cell receptors. Journal of Immunotherapy. 2010;33(6):648–658. doi:10.1097/CJI.0b013e3181e311cb

18. Ito F, Carr A, Svensson H, Yu J, Chang AE, Li Q. Antitumor Reactivity of Anti-CD3/Anti-CD28 Bead-Activated Lymphoid Cells: Implications for Cell Therapy in a Murine Model. Journal of Immunotherapy. 2003;26(3):222–233. doi:10.1097/00002371-200305000-00006

19. Matus EI, Sparkes A, Gariépy J. A soluble activator that favors the ex vivo expansion of CD8+CD27+T cells. JCI Insight. 2020;5(22). doi:10.1172/jci.insight.141293

20. Sadelain M. CAR therapy: The CD19 paradigm. Journal of Clinical Investigation. American Society for Clinical Investigation. 2015;125(9):3392–3400. doi:10.1172/JCI80010

21. Kim GB, Aragon-Sanabria V, Randolph L, et al. High-affinity mutant Interleukin-13 targeted CAR T cells enhance delivery of clickable biodegradable fluorescent nanoparticles to glioblastoma. Bioact Mater. 2020;5(3):624–635. doi:10.1016/j.bioactmat.2020.04.011

22. Ravi K, Trottier S, Amissah OB, et al. Multimodal profiling of CAR T cells against glioblastoma using a microengineered 3D tumor-on-a-chip model. Bioact Mater. 2026;59:724–744. doi:10.1016/j.bioactmat.2026.01.003

23. Lisby AN, Carlson RD, Baybutt TR, Weindorfer M, Snook AE. Evaluation of CAR-T cell cytotoxicity: Real-time impedance-based analysis. In: Spada S, Galluzzi L, eds. Methods in Cell Biology. Vol 167. Academic Press; 2022:81–98. doi:10.1016/bs.mcb.2021.08.002

24. Monfrini C, Stella F, Aragona V, et al. Phenotypic Composition of Commercial Anti-CD19 CAR T Cells Affects In Vivo Expansion and Disease Response in Patients with Large B-cell Lymphoma. Clinical Cancer Research. 2022;28(15):3378–3386. doi:10.1158/1078-0432.CCR-22-0164

25. Shune L, Frigault MJ, Riedell PA. CAR-T cell therapy in older adults with relapsed/refractory LBCL: benefits and challenges. J Immunother Cancer. BMJ Publishing Group. 2025;13(6). doi:10.1136/jitc-2024-009793

26. Schuster SJ, Bishop MR, Tam CS, et al. Tisagenlecleucel in Adult Relapsed or Refractory Diffuse Large B-Cell Lymphoma. New England Journal of Medicine. 2019;380(1):45–56. doi:10.1056/nejmoa1804980

27. Abramson JS, Palomba ML, Gordon LI, et al. Lisocabtagene maraleucel for patients with relapsed or refractory large B-cell lymphomas (TRANSCEND NHL 001): a multicentre seamless design study. Lancet. 2020;396(10254):839–852. doi:10.1016/S0140-6736(20)31366-0

28. Moeller M, Haynes NM, Kershaw MH, et al. Adoptive transfer of gene-engineered CD4+ helper T cells induces potent primary and secondary tumor rejection. Blood. 2005;106(9):2995–3003. doi:10.1182/blood-2004-12-4906

29. Song DG, Powell DJ. Pro-survival signaling via CD27 costimulation drives effective CAR T-cell therapy. Oncoimmunology. Landes Bioscience. 2012;1(4):547–549. doi:10.4161/onci.19458

30. Bai Z, Woodhouse S, Zhao Z, et al. H E A L T H A N D M E D I C I N E Single-Cell Antigen-Specific Landscape of CAR T Infusion Product Identifies Determinants of CD19-Positive Relapse in Patients with ALL. Vol 8. 2022.

31. Chen H, Wei F, Yin M, et al. CD27 enhances the killing effect of CAR T cells targeting trophoblast cell surface antigen 2 in the treatment of solid tumors. Cancer Immunology, Immunotherapy. 2021;70(7):2059–2071. doi:10.1007/s00262-020-02838-8

32. Lee SY, Lee DH, Sun W, et al. CD8 + chimeric antigen receptor T cells manufactured in absence of CD4 + cells exhibit hypofunctional phenotype. J Immunother Cancer. 2023;11(11). doi:10.1136/jitc-2023-007803

33. Xiao X, Huang S, Chen S, et al. Mechanisms of cytokine release syndrome and neurotoxicity of CAR T-cell therapy and associated prevention and management strategies. Journal of Experimental and Clinical Cancer Research. BioMed Central Ltd. 2021;40(1). doi:10.1186/s13046-021-02148-6

34. Manni S, Del Bufalo F, Merli P, et al. Neutralizing IFNγ improves safety without compromising efficacy of CAR-T cell therapy in B-cell malignancies. Nat Commun. 2023;14(1). doi:10.1038/s41467-023-38723-y

35. Zhu X, Li Q, Zhu X. Mechanisms of CAR T cell exhaustion and current counteraction strategies. Front Cell Dev Biol. Frontiers Media S.A. 2022;10. doi:10.3389/fcell.2022.1034257

36. Hammink R, Weiden J, Voerman D, et al. Semiflexible Immunobrushes Induce Enhanced T Cell Activation and Expansion. ACS Appl Mater Interfaces. 2021;13(14):16007–16018. doi:10.1021/acsami.0c21994

37. Jin W, Tamzalit F, Chaudhuri PK, Black CT, Huse M, Kam LC. T cell activation and immune synapse organization respond to the microscale mechanics of structured surfaces. Proc Natl Acad Sci U S A. 2019;116(40):19835–19840. doi:10.1073/pnas.1906986116

38. Casati A, Varghaei-Nahvi A, Feldman SA, et al. Clinical-scale selection and viral transduction of human naïve and central memory CD8+ T cells for adoptive cell therapy of cancer patients. Cancer Immunol Immunother. 2013;62(10):1563–1573. doi:10.1007/s00262-013-1459-x

39. Imam Z, Marsh D, Haines L, Hodge C. EXPANSION OF STEM-LIKE T CELLS BY NOVEL NANOSPARKTM STEM-T ACTIVATOR. Cytotherapy. 2024;26(6):S171–S172. doi:10.1016/j.jcyt.2024.03.336

40. Locke FL, Rossi JM, Neelapu SS, et al. Tumor burden, inflammation, and product attributes determine outcomes of axicabtagene ciloleucel in large B-cell lymphoma. Blood Adv. 2020;4(19):4898–4911. doi:10.1182/BLOODADVANCES.2020002394

