## Supplemental Material for "A soluble bi-specific fusion protein for the improved expansion of human CD8^+^ CAR-T cells"

### Supplemental Methods

#### T-CEP protein expression and purification

The linear bispecific protein T-CEP was produced as previously described.[19] Briefly, a synthetic DNA gene encoding an anti-human CD28-scFv linked to an anti-human CD3-scFv was cloned into a pcDNA-3.4 expression plasmid and used to transform Expi293F cells (ThermoFisher) resulting in the expression and secretion of T-CEP. T-CEP was purified using HisTrap HP columns (GE Healthcare), then desalted into PBS using PD-10 columns (Cytiva). The purified protein was subsequently passed through endotoxin removal columns (ThermoFisher) and sterilized by filtration using a 0.22 μM filter.

#### PBMCs and T cell isolation

Human peripheral blood mononuclear cells (PBMCs) were isolated from EDTA-treated whole blood samples of healthy donors using density gradient centrifugation (Ficoll-Paque Plus, GE Healthcare). PBMCs were cryopreserved in heat-inactivated fetal bovine serum (HI-FBS) (Wisent Inc.) with 10% DMSO (ThermoFisher) in liquid nitrogen until experiments were conducted. PBMCs were collected in accordance with the guidelines set forth by the Institutional Sunnybrook Research Ethics Board under the Project Identification Number 2978. $CD3^+$ T cells were isolated from PBMCs using the EasySep™ Human T Cell Isolation Kit (STEMCELL Technologies) according to manufacturer's instructions.

#### CD19 CAR-T cell generation

Isolated $CD3^+$ human T cells were plated in a flat-bottom 96-well tissue culture plate (Corning Inc.) at a density of $1x10^5$ cells per well in 200 μl of T cell medium in duplicate wells, comprised of X-VIVO 15 Hematopoietic Cell Medium (Lonza) supplemented with 5% HI-FBS and 100 IU/ml of recombinant human IL-2 (animal component free, STEMCELL Technologies). Cells in each well were stimulated with Dynabeads (1:1 bead to cell ratio), human T cell TransAct according to manufacturer's protocols or T-CEP (10ng/ml). Twenty-four hours after T cell activation, 100 µl of T cell medium was removed, and 100 µl of T cell medium containing a lentivirus vector encoding a GFP-tagged CD19-CAR (anti-CD19 scFv-CD8-4-1BB-CD3ζ, eGFP Clone FMC63; BPS Bioscience) was added at a MOI of 20:1. Two days after lentivirus addition, T cells from duplicate wells were combined and transferred to a G-Rex 24-well plate (Wilson Wolf). Dynabeads were removed by magnetic separation and TransAct was removed by washing the cells. The T cell medium containing 100 IU/ml IL-2 was exchanged every three days through replacing 75% of the spent medium. T cell expansion was monitored by counting cells with a haemocytometer. On day 12 of culture, CAR-T cells were sorted by fluorescence activated cell sorting (BD FACSAria) based on GFP expression and DAPI exclusion for viable cells.

**TV-13 CAR Lentivirus generation**

TV-13 was inserted into a second-generation CAR cassette containing a CD8α signal peptide, CD8α hinge and transmembrane domains, a 4-1BB costimulatory intracellular signaling domain (ICD), and a CD3ζ activation domain.[22] GFP-tagged TV-13 CAR lentiviral particles were generated using a previously described protocol.[21,22] Briefly, the TV-13 CAR plasmid was combined with third-generation packaging plasmids pMDLg/pRRE, pRSV-Rev, and pMD2.G in Opti-MEM and incubated for 5 min at room temperature. A separate mixture of Opti-MEM and Lipofectamine 2000 was prepared and incubated for 5 min at room temperature. The plasmid mixture was then added dropwise to the Lipofectamine 2000 mixture to generate the transfection medium and incubated for 20 min at room temperature. The transfection medium was added dropwise to 70–80% confluent HEK293T cells, and cells were incubated at 37°C and 5% $CO_2$. Lentivirus-containing supernatants were collected at 24 and 48 h after transfection, filtered, and concentrated by ultracentrifugation at 25,000 rpm for 2.5 h. Lentiviral particles were harvested, aliquoted, and stored at −80°C until use.

**TV-13 CAR-T cell generation**

Isolated $CD3^+$ T cells were seeded at $1x10^5$ cells per well in a flat-bottom 96-well tissue culture plate (Corning Inc.) in 200 µl of T cell medium. Cells were stimulated with Dynabeads, TransAct or T-CEP (10ng/ml). Twenty-four hours after activation, 100 µl of T cell medium was removed from each well, and 100 µl of fresh pre-warmed T cell medium containing TV-13 lentivirus was added to the cells at an MOI of 20:1. T cell medium (100 µl) without lentivirus was added to a well of activated T cells as a negative, untransduced control. Dynabeads were removed by magnetic separation 48 hours after lentivirus addition and TransAct was removed by centrifugation at 300*x g* for 7 min at room temperature. Cells were resuspended in fresh T cell medium and transferred to G-REX 24-well plates. The spent medium (75%) was replaced with fresh, pre-warmed T cell medium supplemented with 100 IU/ml IL-2 every 3 days for 13 days. Transduction and viability were assessed by flow cytometric analysis of GFP-expression and fixable viability dye exclusion (eBioscience™ Fixable Viability Dye eFluor™ 450, ThermoFisher). Cell expansion was monitored by counting cells using a hemocytometer.

**xCELLigence real time cell analysis (RTCA) cytotoxicity assay**

The xCELLigence RTCA DP system (Agilent Technologies) was used to monitor $CD19^+$ Daudi (CCL-213) cell growth and killing by CD19 CAR-T cells as previously described.[23] Briefly, anti-CD40 antibodies were immobilized on E-Plate 16 (Agilent Technologies) using the xCELLigence immunotherapy IMT assay (anti-CD40) tethering kit (Agilent Technologies). $CD19^+$ Daudi cells were seeded at a density of $4x10^4$ cells in 200 µl of complete RPMI (Wisent Inc.; RPMI supplemented with 10% HI-FBS and 1% Penicillin and Streptomycin). Cell growth was monitored in the xCELLigence system at 37°C, 5% $CO_2$. After 24 hours, the medium in each well was removed along with any non-adhered cells. Sorted CD19 CAR-T cells were seeded onto target Daudi cells in 200 µl of complete RPMI at a 5:1 effector-to-target cell ratio in duplicate or triplicate technical replicates for each donor. As a negative control, 200 µl of fresh complete RPMI was added to wells containing Daudi cells alone. Monitoring was continued for another 40 hours.

To monitor TV-13 CAR-T cell cytolytic activity, IL-13Rα2-expressing target U87 cells were seeded at 5,000 cells per well on E-Plate 16 in 200 µl of complete DMEM (Wisent Inc.; DMEM supplemented with 10% HI-FBS and 1% penicillin and streptomycin). Twenty-four hours after U87 seeding and adherence, complete DMEM was removed, and TV-13 CAR-T cells were seeded at a 5:1 effector-to-target ratio over adherent U87 cells in 200 µl of T cell medium. As negative controls, T cells were seeded in 200 µl of T cell medium without U87 target cells. T cell medium (200 µl) without CAR-T cells was added to adherent U87 cells as an additional negative control. Cells were plated in technical duplicates. Monitoring resumed for another 46 hours.

For both CAR-T cell models, each experiment was performed using two biological replicates, for a total of five to six biological replicates across three independent experiments. Cell Index was recorded as electrical impedance caused by cell adherence using RTCA Software Pro (Agilent Technologies). Normalized Cell Index is calculated as per manufacturer's Calculation Principles protocol using the following equation:

$$\text{Normalized Cell Index}(t_n) = \frac{\text{Cell Index}(t_n)}{\text{Cell Index}(t_0)}$$

where $t_n$ = Cell Index at a given time, and $t_0$ = Cell Index just before the addition of CAR-T cells. Cytotoxic activity is reported as percent cytolysis using the following equation:

$$\%\text{Cytolysis} = \frac{(\text{Cell Index}_{\text{no effector}} - \text{Cell Index}_{\text{effector}})}{\text{Cell Index}_{\text{no effector}}} \times 100$$

**Flow cytometry**

For CD19 CAR-T cells, harvested T cells were incubated with Human TruStain FcX (BioLegend) for 10 minutes on ice to block Fc receptors. The cells were then stained with Alexa Flour 647 anti-human CD8 (RPA-T8; BioLegend), PE/Cy7 anti-human CD4 (OKT4; BioLegend), PE/Dazzle™ 594 anti-human CD27 (0323; BioLegend), and APC/Cy7 anti-human CD45RA (HI100; BioLegend) on ice for 30 minutes in Hanks buffered saline solution (HBSS) (Wisent Inc.). Cells were washed once with HBSS, then resuspended in 30 nM DAPI (ThermoFisher) prepared in PBS.

To monitor the transduction and phenotype of TV-13 CAR-T cells throughout expansion, T cells were incubated with Human TruStain FcX for 10 minutes at 4°C to block Fc receptors. Cells were stained with eBioscience™ Fixable Viability Dye eFluor™ 450 (ThermoFisher), Alexa Flour 647 anti-human CD8 (RPA-T8; BioLegend), PE/Cy7 anti-human CD4 (OKT4), PE/Dazzle™ 594 anti-human CD27 (0323), and APC/Cy7 anti-human CD45RA (HI100) in FACS buffer (PBS without $Ca2^{+}$ and $Mg^{2+}$ supplemented with 2% HI-FBS) at 4°C for 30 minutes. To assess activation, T cells were restimulated with or without U87 cells for 46 hours. T cell medium was removed, and cells were washed once in FACS buffer. Cells were then incubated with Human TruStain FcX for 10 minutes at 4°C to block Fc receptors. T cells were incubated with eBioscience™ Fixable Viability Dye eFluor™ 450, Brilliant Violet 711 anti-human CD8 (SK1; BioLegend), PE/Cy7 anti-human CD4, PE/Dazzle™ 594 anti-human CD27 (0323), APC/Cy7 anti-human CD45RA (HI100), PE anti-human CD25 (OKT4) or PE anti-human PD-1 (EH12.2H7), and APC anti-human CD69 (FN50) or APC anti-human LAG-3 (7H2C65) for 30 minutes at 4°C. Cells were washed twice with FACS buffer, then resuspended in FACS buffer and analyzed on the BD FACSymphony A3 Flow Cytometer.

**Cytokine analysis**

Cytokine secretion was measured to further assess TV-13 CAR-T cell function. Following 46 hours co-culture with or without U87 target cells, cell culture supernatants were harvested into 96-well round-bottom plates (Corning Inc.) and centrifuged twice at 3,000 rpm to remove any debris. Supernatants were stored at -80°C until time of assay. The LEGENDplex™ Human CD8/NK mix and match kit was used to measure the secretion of IFN-γ, TNF-α, IL-4 and Granzyme B (BioLegend). The assay was performed per manufacturer's instructions and samples were analyzed using the BD FACSymphony A3 Flow Cytometer.

**Data analysis**

Flow cytometric analyses were performed using FlowJo v10 (BD Biosciences). Secreted cytokines were analyzed using BioLegend LEGENDplex™ Data Analysis Software Suite fitted with a 5-parameter logistic curve, then normalized by square root transformation. Statistical analyses were performed using GraphPad Prism 10. Results with a $p$-value <0.05 were considered significant.

**Supplemental figures**

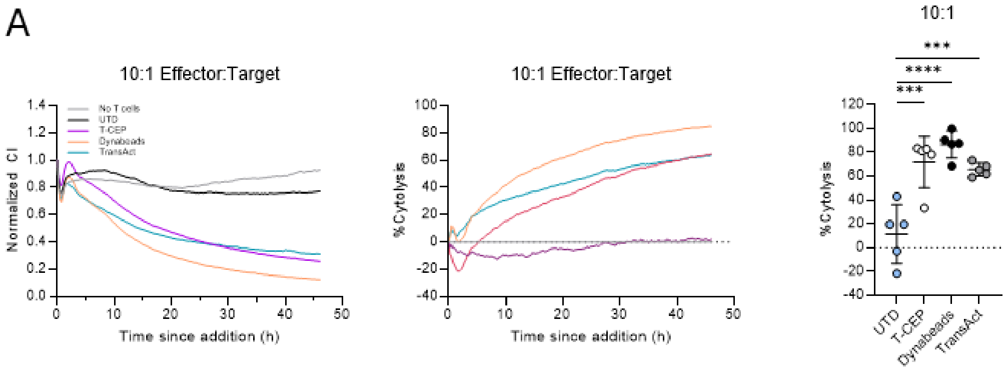


**Supplemental figure 1. TV-13 CAR-T cells killing of IL-13Rα2-expressing U87 cells at 10:1 effector:target ratio.** To further assess CAR-T cell cytotoxicity, CAR-T cells were also co-cultured at a higher E:T ratio of 10:1 for 46h. Plots show Normalized Cell Index (CI) over 46h of co-culture (left), percent cytolysis over time (middle) and statistical analysis of percent cytolysis at after 46h co-culture (right). Left and middle panels show mean, right panel shows mean ± SD, n=5. *** *p<0.001, **** p<0.0001.*

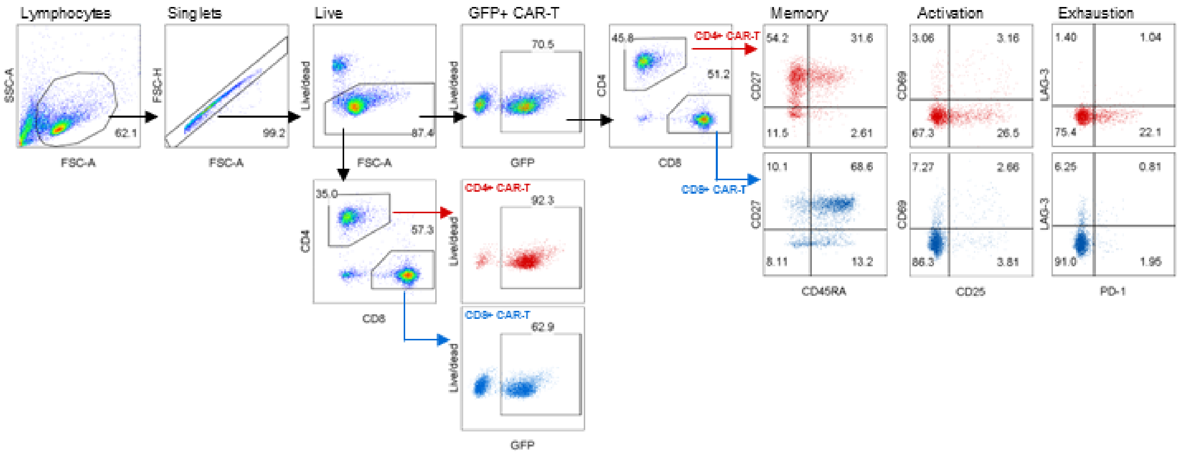


**Supplemental figure 2. Gating strategy.** CAR-T cells were analyzed by flow cytometry after 46h co-culture with U87 target cells at 5:1 effector:target ratio. Cells were first gated for Lymphocytes based on FSC-A and SSC-A, then Singlets based on FSC-H and FSC-A. Live cells were gated based on exclusion of the amine reactive Fixable-Viability dye, eFluor 450. Next, CAR-T cells were identified based on GFP expression, followed by $CD4^+$ or $CD8^+$ expression. The expression of memory markers (CD45RA, CD27), activation markers (CD69, CD25) and exhaustion markers (PD-1, LAG-3) were then analyzed on each subset. To assess transduction efficiency, $CD4^+$ and $CD8^+$ subsets were gated right after dead cell exclusion. GFP expression was then analyzed within live $CD4^+$ or $CD8^+$ T cells.
